# A Generalizable, Governance-Embedded Pathway for Assessing Patient-Level Opportunities for Genetic Medicines

**DOI:** 10.64898/2026.09.22.26363712

**Authors:** David Cheerie, Amy Y. Pan, Meryl Acker, Kimberly Amburgey, Jashanpreet Sidhu, Laura Buckley, Hernan Gonorazky, Nicole K. McKinnon, Lyndsey McRae, Miriam S. Reuter, Lindsey Vogt, Laura Zahavich, Tomasz Czarny, Eriskay Liston, Andrew McFadyen, Daniel A. Morgenstern, Brian T. Kalish, Michelle M. Axford, Ashish R. Deshwar, Zhenya Ivakine, James J. Dowling, Gregory Costain

**Author notes:** Correspondence: Gregory Costain. These authors contributed equally (David Cheerie, Amy Y. Pan, Meryl Acker, Kimberly Amburgey). These authors contributed equally (James J. Dowling, Gregory Costain).

## Abstract

**Purpose:** To describe the development of - and outcomes from - an institution-level initiative supporting access to innovative genetic medicines information and opportunities.

**Methods:** We developed and implemented a Genetic Medicines Patient Assessment Pathway for rare genetic conditions at a large public hospital, with multidisciplinary teams collaborating to evaluate a patient’s molecular and clinical amenability to experimental and/or to-be-developed genetic medicines (primarily nucleic acid therapeutics). A Genetic Medicines Consult Clinic operating in parallel offered no-cost specialized consultations and counseling to families regarding genetic medicines inquiries and Pathway outcomes. We retrospectively reviewed submissions, decisions, and outcomes over a >2-year period.

**Results:** A total of 63 submissions were reviewed in the Patient Assessment Pathway. Most submissions were classified as eligible or potentially eligible for at least one genetic medicines approach (42/63; 67%). Recommendations generated 87 discrete actions, such as outreach to potential collaborators (n = 28), offering participation in existing research studies (n = 23), or referral to the Consult Clinic (n = 19). Two patients have been approved for therapeutic program initiation or clinical trial enrolment.

**Conclusion:** Findings to date support feasibility and high uptake of a structured, multidisciplinary, medical genetics-led model for assessing patient-level opportunities for genetic medicines.

## INTRODUCTION

The therapeutic landscape for genetic conditions is changing quickly, with expanding boundaries of possibility. Advances in genetic medicines, including but not limited to nucleic acid therapeutics like antisense oligonucleotides (ASOs), gene replacement, and gene editing, have created newfound hope and expectations in the rare disease community.^1–4^ Although relatively few genetic conditions have disease-modifying therapies available through the clinic, early-stage clinical trials are increasingly common and there is proof-of-concept for individualized genetic interventions.^5–10^ Clinicians are increasingly asking about - or questioned by patients and families about - the feasibility of an innovative genetic medicines program for a rare or ultra-rare genetic condition.^11^

Supporting patients with confirmed genetic diagnoses and unmet therapeutic needs, including those families on “therapeutic odysseys”, is fundamental in modern medical genetics practice.^11–14^ Research advances in the genetic medicines space present both opportunities and significant challenges for clinicians and their institutions.^11,12,15–17^ At our large, publicly funded, pediatric academic hospital, institutional leadership recognized the need for a transparent, equitable, and evidence-based framework to support decision making around experimental genetic medicines. We aimed to address this gap in medical genetics workflows via new and formalized processes. Here, we describe the implementation of the Genetic Medicines Patient Assessment Pathway (a decision-making framework) and a companion Genetic Medicines Consult Clinic (a patient- and family-facing communication arm), and evaluate referral patterns, activities, and outcomes over a >2-year period.

## METHODS

This is a retrospective, single-center study approved by the Research Ethics Board at The Hospital for Sick Children (SickKids) with an accompanying consent waiver. The Genetic Medicines Patient Assessment Pathway and the Consult Clinic are described below and in Figure

1. Pathway structures and membership responsibilities were codified in Terms of Reference. All information received from case submissions was stored in a secure Microsoft SharePoint list. We extracted non-identifying inquiry submission data from February 2024 to April 2026, and assessment findings and outcomes from the three phases of the Pathway. Non-identifying data were also extracted from clinical records and summarized for the Genetic Medicines Consult Clinic.

### Phase 1: Triage Working Group

Pathway submissions requesting an assessment of one or more specific DNA variants are reviewed within 30 days, during a weekly scheduled meeting. Submissions are made by clinicians and/or members of the care team. Required input is variant-level details, and basic demographic and clinical phenotype data. The goal is to review molecular genetic considerations in relation to current or short-term (1-2 years) future genetic medicine development opportunities, not influenced by patient/family factors or the potential for funding. The Triage Working Group (Figure 1) is comprised of five clinicians and researchers with expertise in rare disease diagnostics and genetic medicines, including two medical geneticists and one molecular genetics laboratory director. The members do not consider, and typically are not aware of, individual patient-level factors beyond basic demographics and clinical phenotypes provided by the submitter. Support staff with scientific backgrounds in genetic medicines prepare dossiers in advance of the meeting that include the following details: (i) pathogenicity evidence for the variants, (ii) evidence supporting the disease-gene association and pathomechanism, (iii) preclinical evidence supporting a therapeutic strategy, (iv) current research opportunities and ongoing therapeutic development efforts.^7,12^ Feedback is returned to all submitters, including specific actionable recommendations for next steps. All submissions were also archived to facilitate periodic re-assessment.

**Figure 1:**
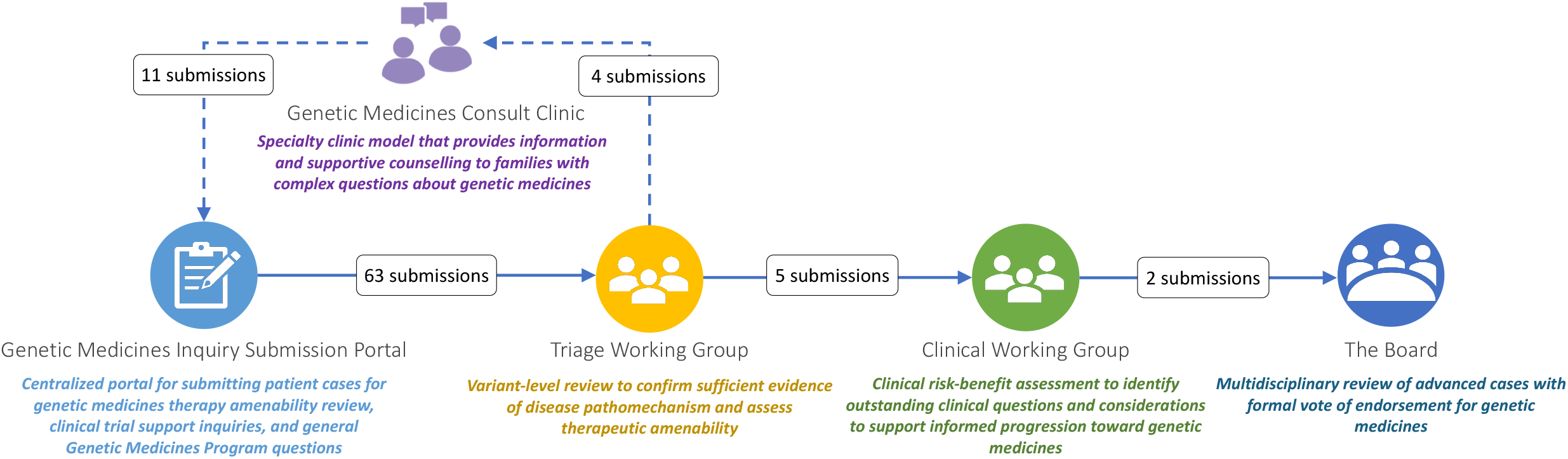
Workflow of the Genetic Medicines Patient Assessment Pathway. Inquiries are submitted through a centralized portal and undergo variant-level review by the Triage Working Group, followed by clinical evaluation by the Clinical Working Group. Selected cases proceed to multidisciplinary Board review. Some cases are referred to, or originate from, the Genetic Medicines Consult Clinic. Numbers indicate submissions at each stage.

Non-mutually exclusive categories of recommendations were: (i) suggesting relevant internal or external research opportunities; (ii) suggesting a referral of the patient to the Genetic Medicines Consult Clinic (described below), and (iii) recommending further review by a Clinical Working Group (described below). Therapeutic eligibility discussion outcomes were categorized as: (i) eligible and actionable (i.e., a genetic medicine targeting the patient’s gene and/or variant is already in clinical trials, irrespective of whether this specific patient would be eligible for the trial), (ii) potentially eligible at this time (i.e., a genetic medicines approach has been studied pre-clinically, or there is a strong precedent genetic medicines for similar diseases and similar variant pathomechanisms), (iii) insufficient information/more research needed (e.g., more evidence needed about variant pathogenicity or disease pathomechanism), and (iv) ineligible at this time (i.e., no existing genetic medicines approach, and no or limited evidence suggesting current genetic medicines approaches are a viable therapeutic strategy).

### Phase 2: Clinical Working Group

Submissions identified as eligible or potentially eligible for a genetic medicines program, and where additional institutional support and investment would be needed to advance such a program, are then discussed in the Clinical Working Group (Figure 1). Meetings occur on an as-needed basis, attended by voting members (5 clinicians, 1 bioethicist), additional ad-hoc members with specific areas of expertise relevant to each case, and representatives from the patient’s clinical teams. This group conducts a structured review of relevant clinical factors, including: (i) clinical history, disease trajectory, and evidence of phenotypic reversibility, (ii) safety, suitability, and risk-benefit considerations, (iii) potential treatment goals, study designs, and clinical outcome measures, (iv) equity considerations and broader applicability of proposed therapy, (v) recommended next steps to support therapeutic development. Voting members of the committee are the clinicians, researchers, and bioethics professionals not directly involved in the patient’s care. The Clinical Working Group either supports the submission moving forward as is to the Genetic Medicines Board or identifies questions in need of further patient/family, clinician, clinical trialist, and/or other researcher input prior to determination of outcome.

### Phase 3: Board

The Board serves as the central decision-making body (Figure 1). Membership includes institutional representatives from clinical departments, research, legal, research ethics, bioethics, and operations, and two external patient family advocates. The Board holistically evaluates the information summarized by the Triage Working Group and Clinical Working Group pertaining to launching or otherwise supporting genetic medicines programs, and votes to either move forward or pause on the proposed plan.

### Genetic Medicines Consult Clinic

The Genetic Medicines Consult Clinic is co-led by a medical geneticist and a genetic counselor, and provides information and supportive counseling to families with complex questions about genetic medicines. Patients were referred either to discuss outcomes of the Patient Assessment Pathway (Figure 1), or by primary care or specialist care providers who determine a family’s questions fall within scope of the clinic. In the latter situation, patients with confirmed genetic diagnoses were eligible when their care teams identified unmet informational or decision-making needs related to genetic medicines. Additional referral criteria, and additional details about clinical flow and structure, are available upon request.

## RESULTS

### Outcomes from Triage Working Group

Overall, 63 submissions were received over the 27-month period (Figure 1). On average, cases were reviewed within 9 days of submission. There were 40 submissions classified as potentially eligible at this time (63%), 18 as ineligible at this time (29%), 3 as insufficient information/more research needed (4.8%), and 2 as eligible and actionable (3.2%) (Figure 2A). Recommendations generated 87 discrete actions across cases (Figure 2A; mean 1.4 actions per submission). Outreach to potential internal or external collaborators was the most frequent recommendation (n = 28), followed by offering participation in existing (typically discovery-focused) research studies (n = 23) or referral to the Genetic Medicines Consult Clinic (n = 19). Five submissions were advanced to the Clinical Working Group.

**Figure 2:**
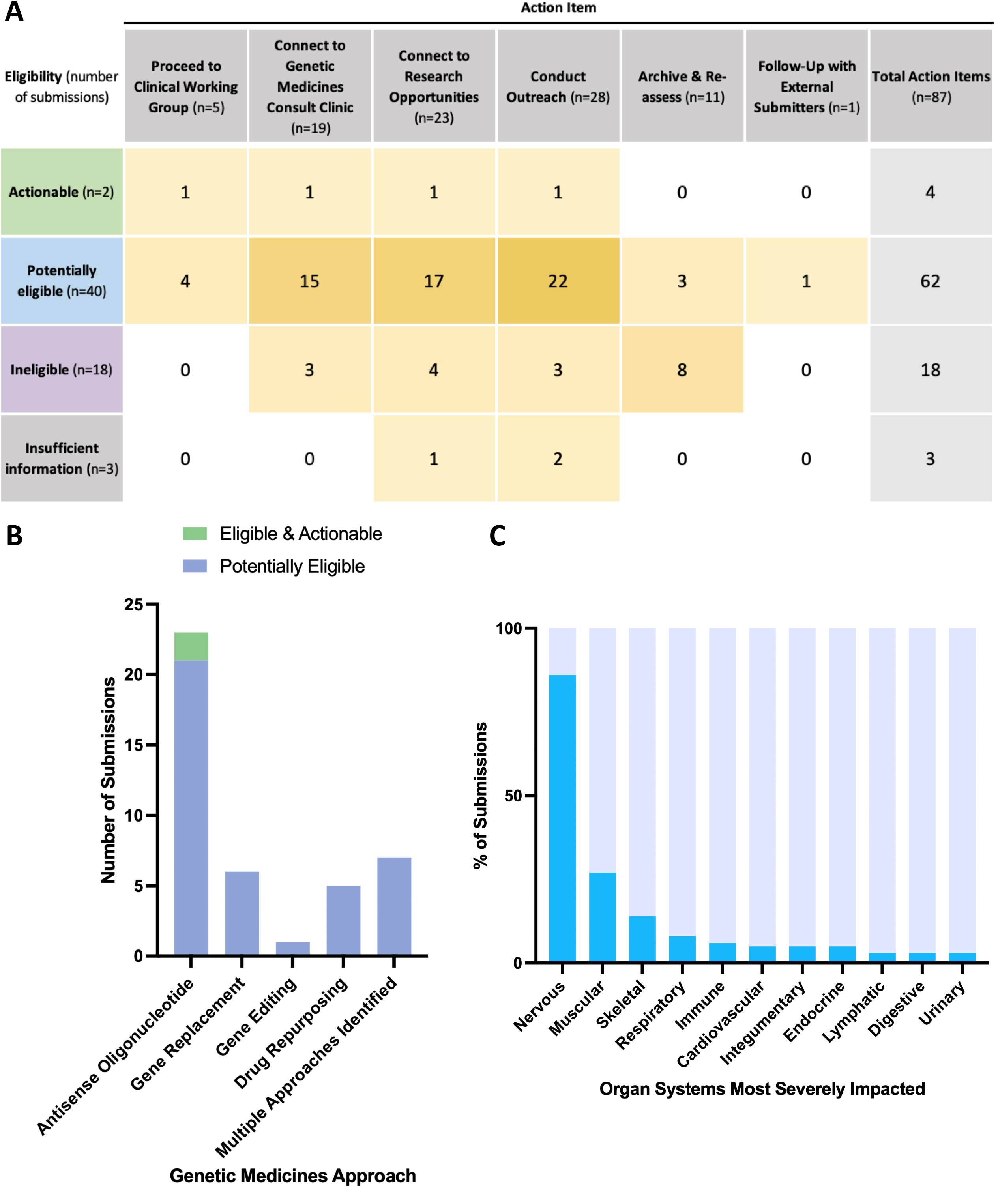
Summary of action items, therapeutic approaches, and organ system involvement for Genetic Medicines Patient Assessment Pathway submissions. **A)** Distribution of action items by eligibility category; most arose from potentially eligible cases and involved outreach, research engagement, and clinic referrals. Eleven cases were archived for re-assessment; one required external follow-up only. **B)** Stacked bar chart of proposed genetic medicines approaches determined to be “eligible and actionable” or “potentially eligible at this time” by the Triage Working Group. **C)** Stacked bar chart of impacted organ systems pertaining to submissions reviewed by the Triage Working Group. As some disorders affect multiple organ systems, these categories are not mutually exclusive.

For both submissions determined to be eligible and actionable, an ASO applicable to the patient’s variant was being evaluated in clinical trials. Of the submissions classified as potentially eligible, 21 involved an ASO approach identified, 6 involved a gene replacement approach, 5 involved a drug repurposing approach, and 1 involved a gene editing approach; the remaining 7 involved multiple possible approaches (two or more of: ASO, gene replacement, gene editing, drug repurposing) (Figure 2B). Most submissions involved disorders affecting the nervous system (54/63 or 86%; including n=2 with retinal disease), the muscular system (17/63 or 27%), and the skeletal system (9/63 or 14%) (Figure 2C). Findings were similar for the subset of submissions identified as actionable or potentially eligible [nervous system (38/42 or 90%); muscular system (11/42 or 26%); immune system (3/42 or 7%)].

### Outcomes from the Clinical Working Group and the Board

To date, five submissions have been reviewed in a Clinical Working Group, including for one disorder predominantly affecting the brain, one retinal disorder, one neuromuscular disorder, one immunological disorder, and one multisystem disorder. The therapeutic modalities under consideration included ASO therapy (n = 2), CRISPR-based gene editing (n = 2; 1 *in vivo* and 1 *ex vivo*), and drug repurposing (n = 1). Two programs have been discussed by the Board and approved for further development and/or clinical trial enrollment, with one pending review as of the data extraction date for this study.

### Metrics from the Genetic Medicines Consult Clinic

Seventeen patients from 16 families were seen in the Consult Clinic. Submissions made on behalf of these families were considered within the Patient Assessment Pathway, except for one individual with a recurrent variant already reviewed in the Triage Working Group for another family. The primary focus of the clinic visits was: review of current state of research related to the diagnosis including therapies being explored (n=10), discussions of research participation and available research opportunities (n=4), and explanations of therapeutic eligibility and treatment limitations (n=2). An additional four submissions reviewed by the Triage Working Group were awaiting visits in the Consult Clinic as of the data extraction date for this study.

## DISCUSSION

We report on the implementation and early outcomes of a generalizable, governance-embedded pathway for assessing patient-level opportunities for genetic medicines based on clinical and scientific merits. This pathway represents a centralized, disease-agnostic, institution-wide approach to identifying and evaluating opportunities for genetic medicines. Our approach is intended to mitigate duplicative therapeutic assessments across programs, and utilizes centralized internal triage to support more efficient use of institutional resources. Portable elements of this approach include the governance structure, variant-level triage, and separation of clinical and decision-making processes.

Our findings demonstrate that timely review of inquiries related to genetic medicines eligibility is feasible within an academic research hospital setting. Over the 27-month period, most submissions were deemed potentially eligible for at least one genetic medicines approach. The phases of assessment offer multiple opportunities for go/no-go decisions. When no immediate therapeutic option or clear path forward is identified, patients and families are redirected toward relevant research opportunities, potential collaborators and champions in their rare disease community, and/or additional opportunities to ask questions (via the Genetic Medicines Consult Clinic). Continuous monitoring and an iterative approach—through ongoing community and multi-disciplinary engagement—will enhance effectiveness of this approach. The Genetic Medicines Consult Clinic represents the only patient-facing component of this pathway. This ensures that communication with patients occurs within a centralized clinical setting, while decision making discussion remains within the clinical and governance teams to support consistent objective evaluation of cases.

This preliminary report is limited by the single-center, retrospective design and the early stage of implementation. We are not yet able to describe long-term clinical impact, and a future consideration is multi-center evaluation to assess generalizability. Few submissions have progressed to therapeutic development amid broader infrastructure, funding, and regulatory barriers. However, this pathway provides meaningful benefit by offering structured guidance, clarity, and support to families navigating complex therapeutic development processes by outlining opportunities for research participation and engagement.

The Patient Assessment Pathway represents only one necessary component of a broader genetic medicines strategy. Within our Genetic Medicines Program, emerging therapeutic technologies and partnership proposals are reviewed by a Technology/Partnership Working Group. Another working group is currently developing equity-informed frameworks to guide prioritization, resourcing, and private pay decisions post-Board review. To meaningfully support access to genetic medicines, institutions must also develop internal capabilities and/or establish external partnerships spanning the entire therapeutic development pipeline, including manufacturing, pre-clinical toxicology, regulatory engagement, and the design and execution of regulated clinical trials.^8,18–20^

We present a model for institutional governance of genetic medicines. Following a systematic, transparent, evidence-informed decision-making framework can enhance equitable access to genetic medicines research opportunities. Medical geneticists are well positioned to lead multidisciplinary efforts at the level of a hospital or institution to respond to the evolving needs of patients with diverse rare genetic conditions, their families, and their other clinicians.

## DATA AVAILABILITY

Identifiable data including unpublished/novel rare variants are not possible to share. All remaining data are included in the main text.

## ACKNOWLEDGEMENTS

We would like to thank our colleagues for their valued contributions to the working groups and initiatives associated with the Genetic Medicines Program: James Anderson, Wendy Bordman, Lauren Dempsey, W. Brent Derry, Nomazulu Dlamini, Carolina Gorodetsky, Michal Inbar-Feigenberg, Christian Marshall, Lucie Perillat, Ori Scott, Ben Shakinovsky, Elizabeth Stephenson, Ana Stosic, Stephanie Telesca

## AUTHOR CONTRIBUTIONS

Conceptualization: J.D., G.C.; Data curation: D.C., A.Y.P., M.A., K.A., J.D., G.C.; Formal analysis: D.C., A.Y.P., M.A., K.A., J.D., G.C.; Funding acquisition: J.D., G.C.; Investigation: D.C., A.Y.P., M.A., K.A., J.D., G.C.; Methodology: D.C., A.Y.P., M.A., K.A., J.D., G.C.; Supervision: J.D., G.C.; Validation: D.C., A.Y.P., M.A. K.A., J.S., L.B., H.G., N.K.M., L.M., M.R., L.V., L.Z., T.C., E.L., A.M., D.A.M., B.T.K., J.D., M.A., A.R.D., Z.I., G.C.; Visualization: G.C., D.C., Writing-original draft: G.C., D.C., A.Y.P., M.A.; Writing-review & editing: D.C., A.Y.P., M.A. K.A., J.S., L.B., H.G., N.K.M., L.M., M.R., L.V., L.Z., T.C., E.L., A.M., D.A.M., B.T.K., J.D., M.A., A.R.D., Z.I., G.C.

## ETHICS DECLARATION

This retrospective cohort study was approved by the SickKids Research Ethics Board (#4232), with a consent waiver.

